# Recovering Mandibular Morphology after Disease with Artificial Intelligence

**DOI:** 10.1101/2020.02.24.20027193

**Authors:** Ye Liang, JingJing Huan, Jia-Da Li, CanHua Jiang, ChangYun Fang, YongGang Liu

**Author notes:** **Corresponding author** Jia-Da Li, School of Life Sciences, Central South University, Changsha, 410078, Hunan Province, China, Tax. **First author** Ye Liang, Department of Stomatology, Xiangya Hospital, Central South University, Changsha, 410008, Hunan Province, China.

## Abstract

Mandibular tumors and radical oral cancer surgery often cause bone dysmorphia and defects. Most patients present with noticeable mandibular deformations, and doctors often have difficulty determining their exact mandibular morphology. In this study, a deep convolutional generative adversarial network (DCGAN) called CTGAN is proposed to complete 3D mandibular cone beam computed tomography (CBCT) data from CT data. After extensive training CTGAN was tested on 6 mandibular tumor cases, resulting in 3D virtual mandibular completion. We found that CTGAN can generate mandibles with different levels and rich morphology, including positional and angular changes and local patterns. The completion results are shown as tomographic images combining generated and natural areas. The 3D generated mandibles have the anatomical morphology of the real mandibles and transition smoothly to the portions without disease, showing that CTGAN constructs mandibles with the expected patient characteristics and is suitable for mandibular morphological completion. The presented modeling principles can be applied to other areas for 3D morphological completion from medical images.

**Clinical trial registration:** This study is not a clinical trial. Patient data were only used for testing in a virtual environment. The use of digital data used in this study was ethically approved.

## Introduction

Some diseases and procedures, such as mandibular trauma and radical surgery for oral cancer, often cause bone dysmorphia and defects^1-4^. In such cases, the exact morphology of the normal mandible before disease is often difficult to obtain because the mandible is already deformed when the doctor first examines it. Additionally, the human mandible has strong specificity, and its morphology varies between individuals^5-9^, making it impossible to use a universal mandibular model as a reference for surgery^10^ and increasing the uncertainty of the surgical effect.

The digital surgical process often requires the expected mandibular reference model^11-14^. In the existing digital surgery process, the common method is to mirror repair or manually look for other similar mandibles for local data fusion and smoothing processing, as shown in Fig. 1(a). A more accurate expected reference model is difficult to achieve, time consuming and difficult to promote in clinical practice. However, rapid routing processing often has poor accuracy. For cumulative bilateral lesions, massive lesions, obvious displacement or lesions cross the middle line, there is still no effective method to predict the expected reference model in clinical practice, as shown in Fig. 1(b).

**Fig. 1.**
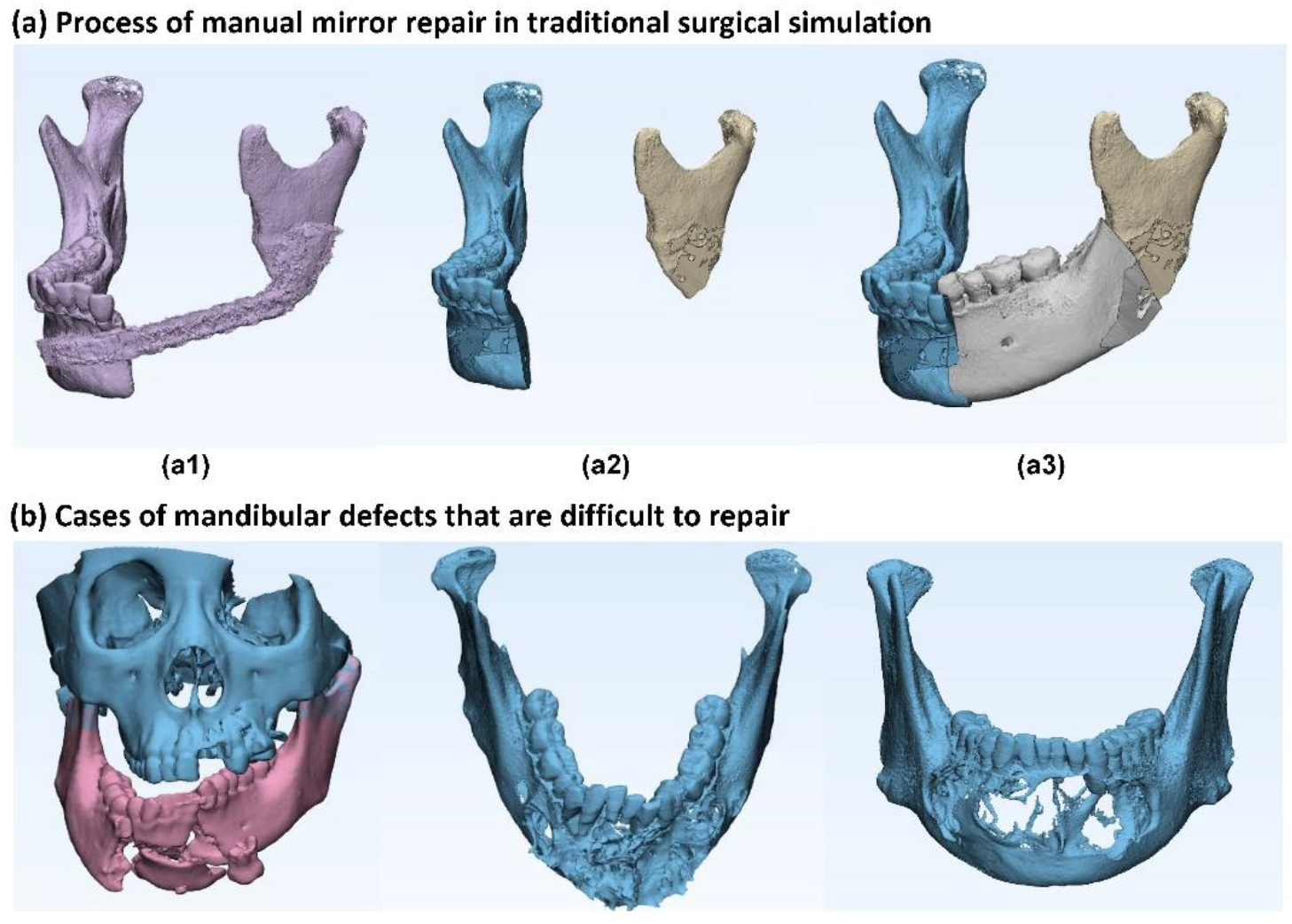
Existing methods and limitations of mandibular reconstruction. In the existing digital surgery process, the common method is to mirror repair or manually look for other similar mandibles for local data fusion and smoothing processing. (**a**) shows the mirror process of manual repair in traditional surgical simulation. (**a1**: original mandible; **a2**: simulation of the removal of all abnormal parts; a3: mirror, crop and splice of the normal parts to achieve the expected reference model; however, the accuracy is often not good). (**b**) shows some cases of mandibular defects that are difficult to repair.

The irreplaceable nature of AI will have important future implications^15-19^. In the medical field, AI technology has also been essential in disease diagnosis and classification, disease image recognition and other fields. Generative adversarial networks (GANs), which endow AI with the capacity to create^20^, were proposed in 2014^21^ and represent the most recent technological breakthrough in the AI field. GANs contain a generative neural network for generating data and a discriminating neural network for determining which data are true. Through unsupervised learning, the two neural networks compete to improve the reliability of the generated results.

This study applies GAN in the mandible medical morphology field. We hope to solve the expected reference model of the mandible, which conforms to both the morphology of the natural mandible and the healthy portion of the patient’s mandibular morphology.

The technique that we applied relies on the deep convolutional generative adversarial network (DCGAN) proposed by Radford et al.^22^ in 2015. This model introduces the GAN algorithm to the random generation of images. The DCGAN learns the morphological features in an image and then uses those features to create entirely new photorealistic images^24,25^. This study used DCGAN to perform machine learning on CT images of normal mandibles to generate images that conform to the anatomical features of the mandible.

Although DCGAN can creatively generate reasonable images that are consistent with the target theme, the resulting images are completely random. With regard to our goal, the generated images conform to only the anatomical mandibular morphology and do not match the patient’s individual morphology. In 2016, Yeh et al.^26^ defined context loss and perception loss based on the DCGAN algorithm and proposed an image semantic repair method for plane images. By iterating randomly generated images, the algorithm continuously reduces the semantic and perceptual losses and finally generates reasonable images that are as similar as possible to the original image in the missing area^27^.

In this study, we aimed to apply DCGAN to CT data, creating CTGAN. We trained the algorithm extensively and tested it in the virtual environment. In this environment we tested 6 mandibular tumors cases, resulting in 3D virtual mandibular completion.

## Results

### Training results of the mandibular discriminator and generator

The goals of the training were to obtain a discriminator that could tell whether the drawing looked similar to a real mandible and to obtain a generator that could draw different tomographic images of the mandible. The discriminator and generator use a randomly initialized neural network. Therefore, their losses change during the training process. A discriminator loss value close to 0 indicates that the discriminator can distinguish the real image from an image generated by the generator. A generator loss value close to 0 means that the image generated by the generator is more likely to be treated as a real image by the discriminator, indicating a high degree of authenticity of the generated image. In Fig. 2, we recorded the loss changes of the discriminator and generator during the 45-hour training (a-b) and clearly observed that the two neural networks improved via the adversarial process (d-e). Fig. 2(c) shows the images generated by the generator with the same input parameters at different training times. More training details of the continuous process are provided in the article video (http://CTGANs.kuye.cn/).

**Fig. 2.**
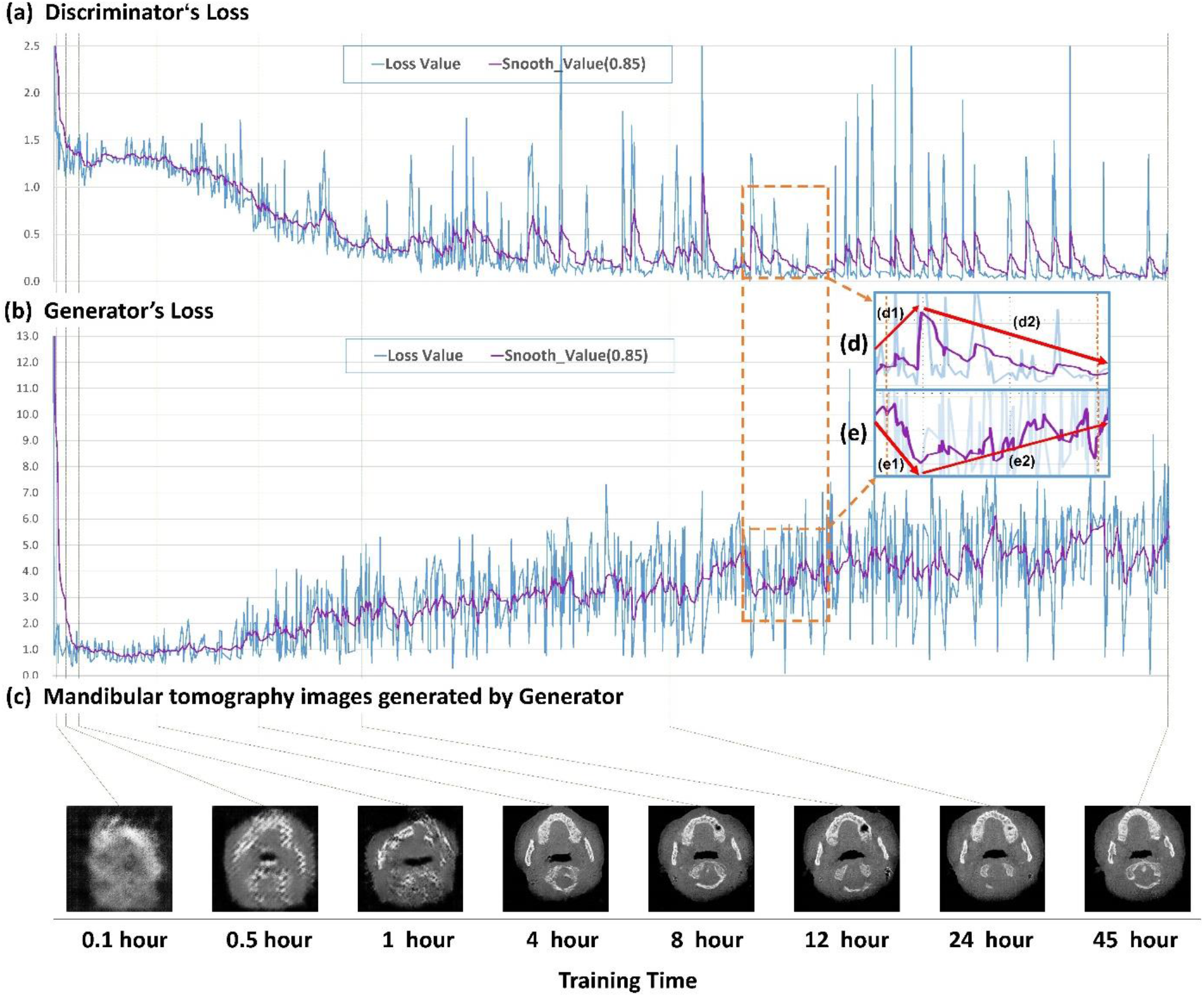
Mandibular tomography generated with different training times. The training time starts with the first batch of mandibular tomography images entering the learning stage. Since the discriminator neural network and the generator neural network are constantly changing during training, the generated image will change. The goal of the training is to obtain a discriminator that can tell whether a drawing looks similar to the mandible and a generator that can draw different tomographic images of the mandible. (**a**) shows the loss value and smooth loss of the discriminator at different training times. A discriminator loss value closer to 0 indicates that the discriminator can distinguish the real image and the image generated by the generator. Since the change in loss value is drastic, we draw the smooth loss with a smoothness of 0.85 to facilitate the analysis. (**b**) shows the loss value and smooth loss of the generator at different training times. A generator loss value closer to 0 means that the image generated by the generator is more likely to be treated as a real image by the discriminator, indicating a high degree of authenticity of the generated image. (**c**) shows the images generated by the generator with the same input parameters at different training times. (**d**-**e**) show the discriminator and generator adversarial training process. In process e1, the loss of the generator decreases, indicating that the generator is making progress, which causes the loss of the discriminator to increase in process d1, in other words, its capacity to judge truth and falsehood decreases. In process d2, the loss of the discriminator decreases, indicating that the discriminator has improved and increasing the loss of the generator. Thus, the authenticity of the images generated by generator needs further improvement. More training details of the continuous process are provided in the article video (http://CTGANs.kuye.cn/).

To determine the performance of the mandibular generator and analyze its robustness, Fig. 3 shows the tomographic image changes in the mandible obtained by regularly adjusting some parameters. We observed the image rotation angle change from 7.0° to -2° and the image position from 0 mm to 9.6 mm. The image local pattern changes with different combinations, and the image layer is also changed. All the changes are continuous. The input parameter of the generator is composed of 100 separate floating-point numbers, and the number of combinations is essentially infinite. With our model, possible mandibular tomographic images can be generated, exceeding the total number of mandibular CT images resulting from all humans on the earth being scanned.

**Fig. 3.**
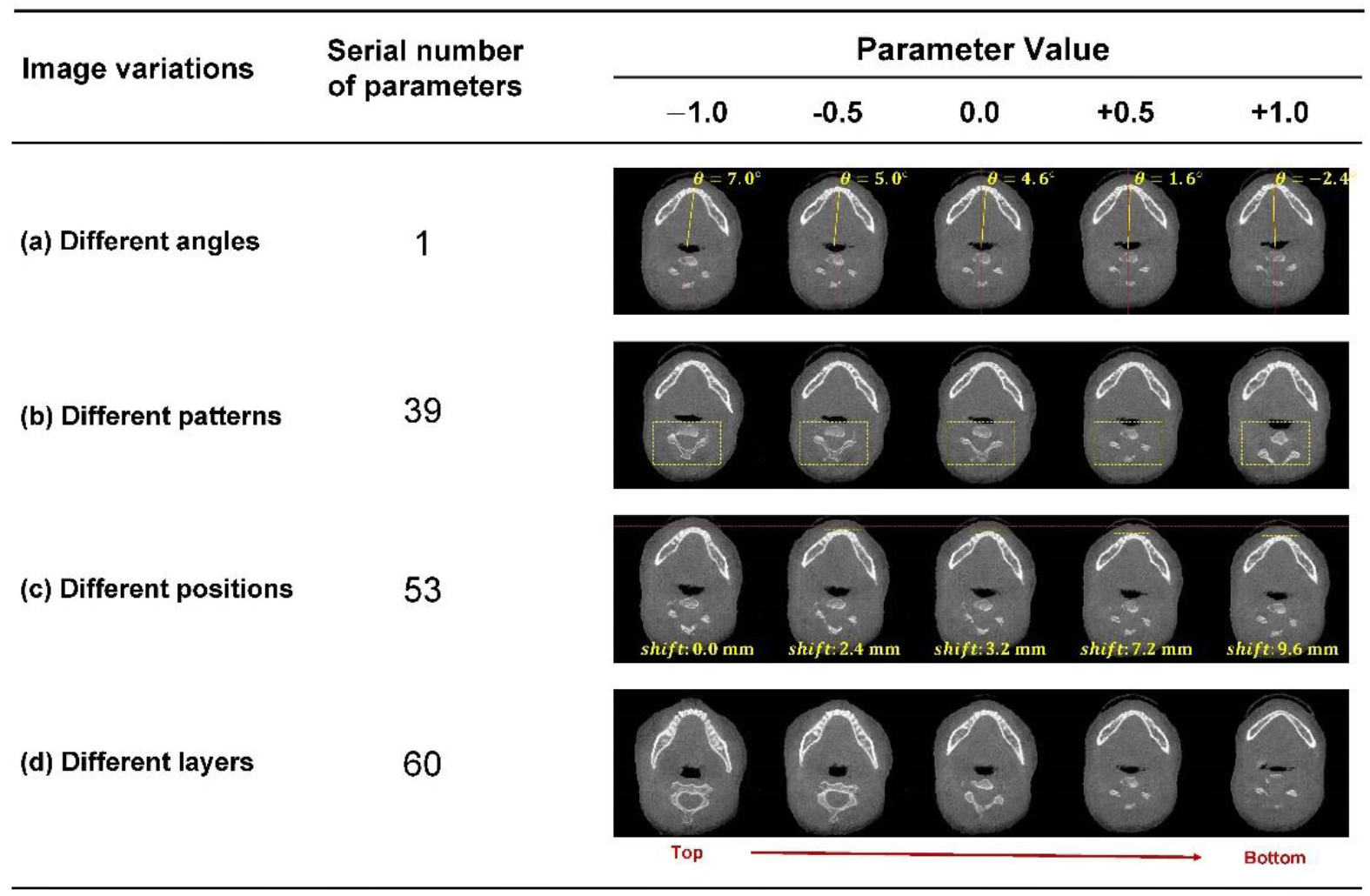
Effect of changing the input parameters on the generated results. The generator’s input parameter is 100 float numbers, and the value range of each number is -1 to +1. We show the results of a sample of random parameters and parameters adjusted using a single variable method. Some changes can be observed in changing parameters of a particular serial number. We list four changes, namely, (**a**) image rotation angle changes (reference, mandibular midline), (**b**) image local pattern changes, (**c**) image position changes (reference, front of mandible), and (**d**) image layer changes. In (**a**-**c**), the changes in the mandibular tomography are measured and marked.

Since the completion model starts with random parameters, the process of obtaining the morphology of a given mandible finds a local optimal solution. In the initial results although many of the completion results were good or perfect, as shown in Fig. 4(a), but some were failures, as shown in Fig. 4(b). As the process is affected by random factors, the incidence rate of failure is between 3% and 57%.

**Fig. 4.**
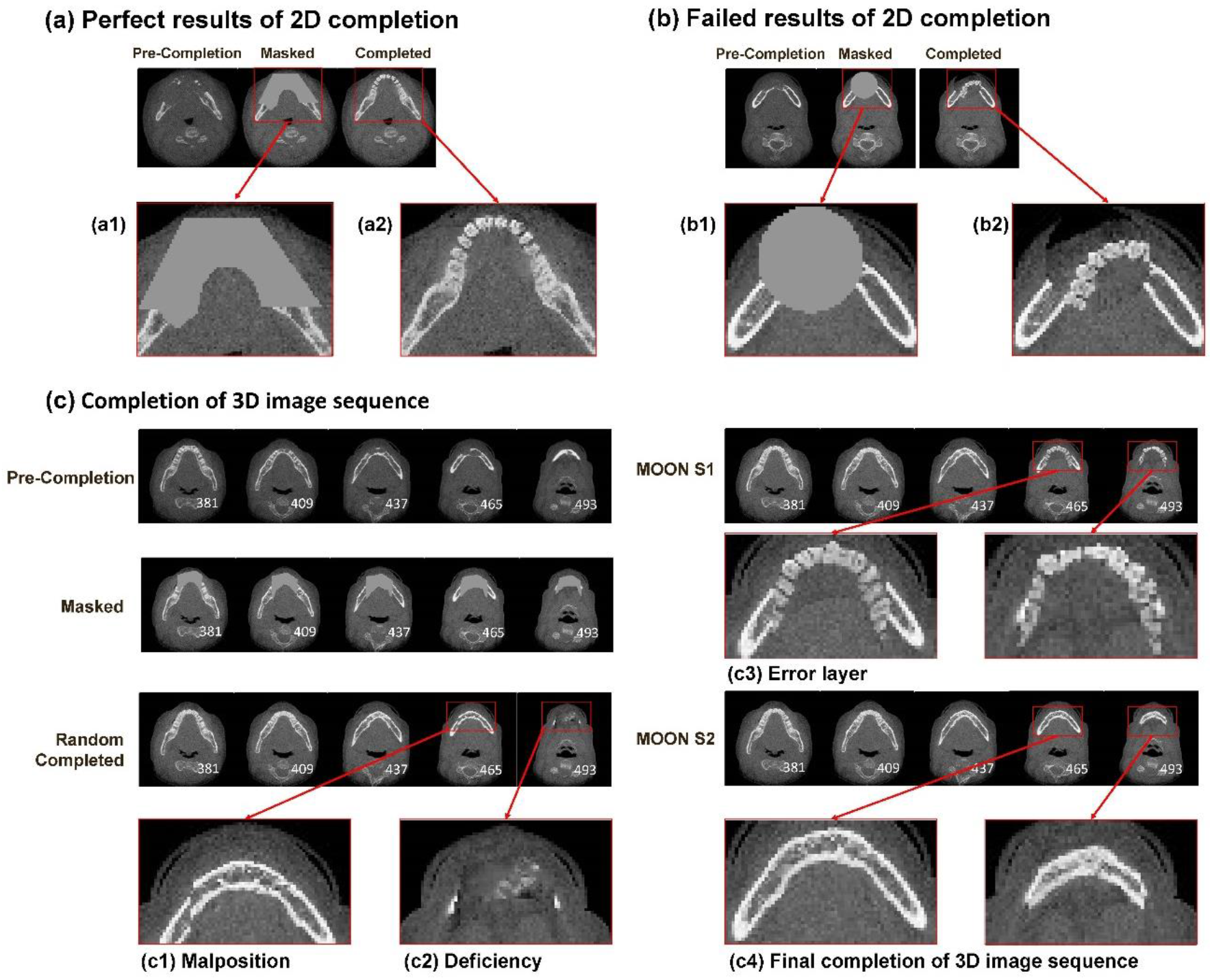
Mandibular tomography completion results. The 2D results of the mandibular tomography completion were extracted and are presented as the perfect result (**a**) and the failed result (**b**). Each set of images contains the precompletion image, the image removal of the mask area, and the completed image. In (**a1**), we remove the lesion area, and we obtain the completion in (**a2**). (**a2**) shows that the completion can combine the generated area with the natural area. Therefore, although many of the completion results were good or perfect, some obviously failed, for example, (**b1**) and (**b2**). (**c**) Shows the completion of the 3D image sequence for images from a real case. Due to the excessive number of images, only one of every 28 images in the lesion area is displayed. First, we use 2D completion initialized with a random input. Although only a few images fail, it is difficult to obtain all perfect results due to the many layers of 3D images. (**c1**) shows a type of failure that is to complete the pattern with a malposition of the natural bone. (**c2**) shows another type of failure, in which part of the jawbone is deficient due to completion. To discard the poor results generated in the completion process, we establish the MOON model. In the first step of MOON, we generate multiple completion results and remove the failed results using the image loss value. However, we still find some mistakes, such as (**c3**). Although the image looks similar to mandibular tomography, it should not have a tooth image in the lower part of the mandible, i.e., the layer is wrong. In the second step of MOON, we calculate the semantic loss of each candidate image of the current tomography and its adjacent layer and obtain the final completed image.

To solve this problem, we added the MOON model for CTGAN. There are many tomographic images in the 3D CT data. Therefore, not all images are completed perfectly, as shown in Fig. 4(c), (c1) and (c2) show some examples of failed 2D completions. In Moon S1, the failed results were excluded by the loss. However, some problems were observed in Fig. 4(c3).

Although the completion has a low loss, in the lower part of mandible, there should not be a tooth; therefore, this layer is an error. To obtain the correct layer and generated mandibular tomography with respect to the spatial correlation of the CT 3D data, we use Moon S2. Fig. 4(c4) shows the final completed image filtered by MOON.

### 3D morphology of the completed mandible

Compared with 2D completion, the most important change in CTGAN is the completion of 3D spatial data. The results of the diseased mandible reconstruction are displayed in 3D. Fig. 5 shows the 3D completion process for 6 cases. One case with a large range defect of the mandible is presented in detail; however, the completed mandible is shown for all cases. Furthermore, we provide additional 3D completion details in the video of this article (http://ctgans.kuye.cn/).

**Fig. 5.**
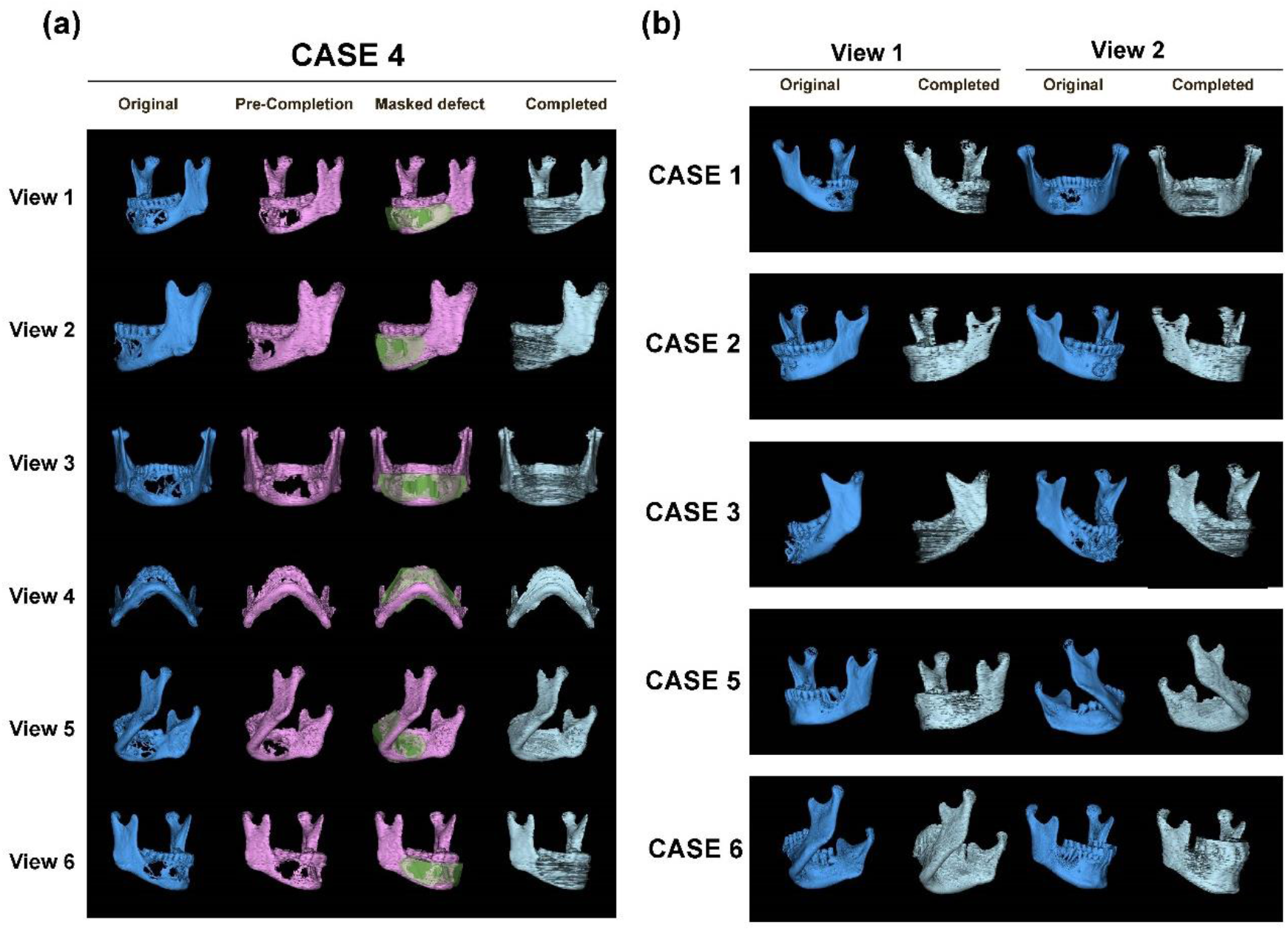
3D mandibular completion in 6 clinical cases. (**a**) shows Case 4, which includes a large range of mandible defects that are difficult to repair using conventional methods. We present the case in detail from six views. The original mandibular morphology from CBCT, the precompletion with reduced resolution, the area to be filled with the 3D mask and the final completion results are shown. (**b**) shows five other cases. For these cases, the completion process was the same as that in Case 4. Therefore, only the two views of the original mandible and the final completion are shown.

## Discussion

Although current medical science cannot provide a mandibular prosthesis for the patient directly from the computer design, the design is significant because it provides a reference for the doctor. In the functional reconstruction of the mandible using fibula or ilium, the reference from mandible completion can help ensure a more accurate splicing position and angle.

The CTGAN model reduces the manual process and improves the mandibular completion efficiency compared with traditional methods such as mirror inversion. The CTGAN model is simple and effective when the lesion crosses the mandible midline. For cases with a wide range of lesions, the CTGAN model completed very little normal morphology information.

In the field of biomaterials research, personalized bone guidance materials are approaching maturity^4,28^; however, obtaining a patient’s personalized morphology is problematic. We believe that the CTGAN model can be used to obtain the natural morphology of the patient’s mandible. In this study, E-3D software for 3D visualization was shown to quickly generate STL files for 3D printing.

According to Fig. 2, the generator was able to capture the initial features of the mandible and create low-quality mandibular images after four hours of training. However, creating high-quality images was more difficult; quality improvement was very slow over the next 4 to 24 hours. Finally, after 45 hours of training, we obtained the generator model.

Generator robustness is mainly determined by the quality and diversity of the generated patterns. All the patterns we observed were high quality. In Fig. 3, we found that parameter changes could not only continuously adjust the generated mandible layer but also affect the image rotation angle and displacement. This finding suggests that during the learning process, the machine automatically correlates two adjacent planes of the mandibular tomography in different orientations or positions and “understands” the reasonableness of the presence in the picture of these types of position and angle changes. Since the argument is a floating point number, the number of patterns that can be generated are theoretically infinite. In addition, we observed that some parameters determine the changes in local patterns in the picture, such as the same layer of the mandibular tomography, which can be matched with different spine morphologies. This suggests that the machine creates images by splitting the CT scan into subpatterns and “understanding” the reasonableness of subpattern matching, which prompts the generator to create more patterns.

Because of the robust generator, we obtain some completion results that can combine the generated area with the natural area. It is difficult to recognize the filling position with professional doctors (Fig. 4a). However, it is also possible to obtain failed completion results that are too poor to be accepted (Fig. 4b). To discard the poor results generated in the completion process, we established the MOON model in CTGAN. MOON can remove failed results from the mandibular completion process and look for context loss from the adjacent layers to seek spatial data continuity. In Fig. 4c, we verified that MOON was actually effective in removing the failed results.

Drawing a 3D model of the completed mandible is an exciting process. In Fig. 5a, the original CBCT mandible, the precompletion with reduced resolution, the area to be filled with the mask and the final completion result are shown in accordance with the completion process detailed in case 4. We use case 4 to present details because of the large mandibular defect across the midline, which leaves little normal information in the tomography, making it difficult to repair using conventional methods. After completion, we obtained better results than expected.

The results showed that not only was the mandibular morphology natural, but the transition to the retained portion was smooth. The 3D completion results of other cases in this study also essentially restored the mandibular morphology, which is shown in Fig. 5b.

## Conclusion

Therefore, it is reasonable to believe that after further optimization, CTGAN is practical. They will greatly shorten the time and technical difficulty involved in mandibular completion and even achieve better results than traditional repair.

More importantly, the CTGAN model proposed in this study has dual naturalization and individualization properties. The success of this technique in 3D mandible generation means that this technique can be used not only for mandibles but also that it is likely to be applicable to other areas of the medical field that involve prosthesis repair.

## Methods

### Ethics statement

This project was approved by the Center of Medical Ethics of Central South University. This study conforms to the Declaration of Helsinki. The training sample was added to the study after removing all of the privacy data. The written informed consent test sample was obtained from the patients. The protection details of the training samples and test samples can be found in the supplementary documentation.

### Dependent work of CTGAN

The DCGAN algorithm and image semantic repair theory based on perception and context loss are mature. A large number of studies have verified and studied DCGAN^23,26,29,30^. Establishing the discriminator and generator in this paper relies on the previous theories of Radford et al.^22^ and Yeh et al.^26,27^, and the main forms of the two networks are the same as those described in their papers, and detail in the Supplementary File. Both the generator and the discriminator are trained using the backpropagation algorithm. The generator finally implements a random generation of the pattern. The loss value generated by the discriminator is used to judge the accuracy of the generated pattern.

### Overview of CTGAN

Since CT data consist of a multilayer image with spatial information, the areas to be completed in each layer are different. Therefore, we optimize the original model and add the definition method of spatial information to be completed. The masking layer is defined three-dimensionally, as shown in the section on establishing mandible test data.

Traditional image completion focuses on only the surrounding information of 2D space, but the context of CT data completion also includes the 3D environment between different layers. If the original algorithm is directly extended to three dimensions, it will be difficult to calculate due to the large quantity of data calculation, or the model cannot be used due to low data precision.

We made a filter using enumeration and dynamic programming algorithms. After the completed results were obtained, they were selected according to the adjacent layer images. The results were better than those without improvement in the preliminary experiment. We describe the results in the optimization of mandible completion results section.

Since this method is optimized for DCGAN using CT data, it is called CTGAN.

### Establishment of mandibular training dataset and test dataset

After preliminary experiments to quickly calculate the neural network convergence, 14,278 images of mandibular tomography were used for neural network training. Among the images, 7392 were from males, and 6886 were from females. We built a training dataset called mini-DICOM. The details are in the supplementary documentation.

A case requiring mandibular completion was the test case for this study. Six patients with mandibular disease who were admitted to Xiangya Hospital of Central South University from March 2018 to February 2019 and voluntarily agreed to undergo digital technology with informed consent were screened. The data were anonymized before using machine learning. In all cases, the mandibular morphology showed obvious changes. In the anterior-posterior, medial-lateral and superior-inferior directions, the maximum lesion distances in the case were 52.89 mm, 70.97 mm and 35.27 mm, respectively. In four cases, the lesion crossed the midline of the mandible; in 5 cases, CBCT was performed in an open position to facilitate the separation of the mandible. More case details are shown in Table 1.

**Table 1.** Basic information used in mandibular completion.

| ID | Diagnosis | Size of Defect (mm) |  |  | Defect<br>Crosses<br>Midline | Shooting Position |
| --- | --- | --- | --- | --- | --- | --- |
|  |  | Bone Axis <sup>a</sup> | Buccal-Lingual <sup>a</sup> | superior-inferior <sup>a</sup> |  |  |
| 1 | ameloblastoma | 39.20 | 12.24 | 25.51 | Yes | mouth opening |
| 2 | ameloblastoma | 37.01 | 12.43 | 27.74 | Yes | mouth opening |
| 3 | ameloblastoma | 129.31 | 28.90 | 30.50 | Yes | mouth opening |
| 4 | ameloblastoma | 95.20 | 27.91 | 33.24 | Yes | mouth opening |
| 5 | left lower<br>gingival cancer | 30.11 | 16.80 | 23.05 | No | mouth opening |
| 6 | right lower<br>gingival cancer | 42.57 | 21.08 | 34.38 | No | mouth closing |

<sup>a</sup> Recorded by the maximum distance of the defect in the corresponding direction.

In the test case, we needed to match the mask for each layer of the images. The mask positions were completed with 0 and the places to be reserved with 1. The size and shape of the mask were consistent with the shape of the CT 3D image. The detailed 3D mask manufacturing method is in the supplementary documentation.

### Training of the mandibular generator

The generation and completion models of this study were completed on the graphics processing unit (GPU) server of the high-performance computing platform of Central South University. The server configuration is shown in the supplementary document.

In this study, CTGAN generators were trained to generate random mandibular images. We trained the mini-DICOM training library with the generator for 260 epochs, which required 45 hours.

To analyze the generator training quality, the results section shows that the generator used ten fixed input parameters, which were produced randomly to generate pictures of the mandible tomography in different training times.

In supplementary document 2, we provide the discriminator and generator loss in the training process.

### Robust analysis of mandibular formation

To determine whether the mandibular morphology generated by the generator is sufficiently diverse and can generate more than learned samples, we adjusted the parameters to assess the robustness of the generator.

is the generator input parameter, formed by 100 independent digits from ‘-1’ to ‘1’. A single variable method was used in the analysis process.

First, a base image was randomly selected to obtain its 100 parameters. Then, we proceeded in turn to change 1 parameter and fix the remaining 99 parameters. The parameter we changed regularly increased.

Changing one of the parameters provided a series of images. The robustness of the generator was judged by a sudden or gradual change in the generated images. The images also helped to infer what the generator had learned.

### 2D evaluation of mandibular completion results

The mandibular completion model was initialized randomly. Therefore, although a pattern similar to the target was obtained by reducing the semantic and perceptual loss as much as possible, the model may still yield a perfect performance or a failed performance. In this section, we show some of the perfect or failed results after completing the mandibular tomography.

### Optimization of mandibular completion results

Compared with 2D image completion, 3D image completion is more difficult to achieve.

In each case, the partial layer image results will fail.

To solve this problem, we set up a multioutcome option network (MOON) in the CTGAN algorithm. MOON used the enumeration method to complete each mandibular tomogram 20-30 times. Failure did not occur every time. MOON first filtered failed results based on the loss value and then calculated the semantic loss of each candidate image of the current tomography and its adjacent layer, forming an undirected graph network. Finally, MOON obtained a set of results with the minimum overall loss through dynamic programming. MOON also allows interactive selection. The addition of MOON achieved better results after some key positions were made clear artificially.

### 3D visualization of mandibular completion

After determining the mandibular completion image of each tomogram, an E-3D digital medical design system can be used for 3D visualization and solid modeling. We show the results of the 3D completion of each case in the Results.

### Data availability

For security and patient privacy reasons, all research data except sensitive information is public. Existing and future research data will continue to be published at http://CTGANs.kuye.cn

## Data Availability

The research data used to support the findings of this study are available from the corresponding author upon request. Training data for some artificial intelligence networks can be downloaded via the data available link.

http://CTGANs.kuye.cn

## Acknowledgements

We are grateful to the High Performance Computing Center of Central South University for assistance with the computations. And thanks to You Zou, Qing Wu for their help in debugging high-performance platforms.

We thank Professor ShengHui Liao (Digital Medicine and 3D Print Research Center, Central South University) for providing the E-3D software and performing targeted optimization of the software. We thank several anonymous reviewers for providing helpful comments on this manuscript. We also thank MAIGO members Jingjing Huan, Kewa Gao, Dan Xu and Shang Fu for their deduction analyses of the mathematical theory. We thank Binzhu Wang, Shouhui Huang, Shanshan Yuan, Yong Zhang, Zhi Chen, and Jin Ai for their assistance with normal mandible CBCT data collection.

This work was financially supported by grants from the National Natural Science Foundation of China (Grant No. 81901065), Hunan Province Science and Technology Department (Natural Science Fund of Hunan Province, 2018JJ3850; Key R&D Program of Hunan Province, 2017SK2161), Health and Family Planning Commission of Hunan Province (20191044), and National Key R&D Program of China (2016YFC1102901).

## Author contributions

YL and JDL raised the concept of the study; YL, JJH, JDL and CHJ designed the study; YL, JJH, CHJ, CYF and YGL collected the data; YL, JDL, CHJ and CYF participated in data interpretation and analysis; YL and JJH prepared the figures; YL, JJH and YGL prepared and edited the manuscript; All the authors reviewed the manuscript.

## Competing interests

The author(s) declare that there are no competing interests.

**Video captions**

This article comes with a Video Summery to quickly explain the content of the article, and more intuitive and concrete results. The video consists of 3 steps. Step 1 shows the training process of the generator. The 9 images in the video are samples from real training. This is the result of the generator generation of 9 constant input parameters during the 0-45 hour training process. This shows the progress that the generator has made in training. In the latter part of the step 1, the background image shows the other generation results using the trained generator in the case of random input, showing the diversity of the generated results. Step 2 shows the 2D image completion process, first marking the lesion and then using the CTGAN algorithm to complete.

In this step, the completion of a CBCT continuous layer in a virtual environment is described in a specific clinical case. Step 3 shows the 3D image completion process. This mandible comes from a patient whose mandibular defect spans the midline and has a large area. First reduce the resolution of the CBCT data and then create a 3D Mask marker defect area. The 3D completion of the jaw was completed by an algorithm. In this step, the original 3D model of the CBCT in this case is given, and the multi-angle topography of the low-resolution model and the complemented model before completion is given.

